# HIF1alpha Cardioprotection in COVID-19 Patients

**DOI:** 10.1101/2021.08.05.21258160

**Authors:** Bingyan J. Wang, Sangeetha Vadakke-Madathil, Lori B. Croft, Rachel I. Brody, Hina W. Chaudhry

**Author notes:** Corresponding author: Cardiovascular Regenerative Medicine, Icahn School of Medicine at Mount Sinai, One Gustave Levy Place.

## Abstract

**Importance:** SARS-CoV-2 infection directly causes severe acute respiratory illness, leading to systemic tissue hypoxia and ischemia including the heart. Myocardial cytopathy associated with hypoxic response has been largely overlooked in COVID-19 patients. Additionally, histology analysis and cardiac function of COVID-19 cases are often reported separately, rendering an incomplete understanding of COVID-19 cardiac symptoms.

**Objective:** To examine the relationship between myocardial cellular responses to hypoxic stress versus cardiac functional alterations within the same COVID-19 patients.

**Design, Setting, and Participants:** Cellular hypoxia Inducible Factor 1 alpha (HIF1α) expression was analyzed by immunohistochemistry using post-mortem COVID-19 heart and lung tissues with known cardiac echocardiography records from a total of 8 patients. Clinical echocardiography data were obtained from Mount Sinai Heart between March to December, 2020. All gender and age groups were considered as long as cardiac involvement meets the preserved (EF > 50%) or moderate to severe (EF < 45%) criteria with confirmed SARS-CoV-2 infection. Cell-type specific subcellular localization of HIF1α expression and nuclear stability was examined by immunohistochemistry and transmission electronic microscopy (TEM). Terminal deoxynucleotidyl transferase dUTP nick end labeling (TUNEL) was used to quantify apoptosis.

**Main Outcomes and Measures:** No planned outcomes of this study as this is a retrospective analysis based on post-mortem specimens exclusively.

**Results:** Cardiac HIF1α expression was found to be significantly higher in patients with preserved EF levels than it was in the low EF group. In the preserved EF group, HIF1α is protective against apoptosis predominantly in endothelial cells and cardiac fibroblasts. In the low EF group, HIF1α protects cardiomyocyte nuclear integrity as evident by its nuclear accumulation with nuclear envelope preservation.

**Conclusions and Relevance:** This study establishes a direct link of cardiac cellular responses to hypoxic stress with matching functional and histological data, serving as one of the first studies to bridge previous stand-alone clinical data and cellular data. The protective role of HIF1α in hearts may help predict cardiac involvement in not only COVID-19 patients, but also decipher the underlying mechanisms in other forms of viral cardiomyopathy.

**KEY POINTS:** *Question:* Are hypoxic signaling pathways associated with cardiac functional alterations in COVID-19 patients?

*Findings:* Cardiac HIF1α expression of COVID-19 patients with EF>50% or EF<45% was analyzed and quantified. Increased cardiac HIF1α^+^ cells were found in patients with higher EF. HIF1α^+^ endothelial cells are resistant to apoptosis, and HIF1α^+^ cardiomyocytes are able to retain nuclear envelope under hypoxic stress.

*Meaning:* HIF1α is cardioprotective in hearts of COVID-19 patients.

## INTRODUCTION

The COVID-19 pandemic has become the most urgent calling of global scientists since it first began in late 2019/early 2020. This respiratory infection has been reported to cause cardiac dysfunction in 25%-55% patients^1,2^, not exclusively in hospitalized patients but also involving patients recovering from non-critical illness^3,4^. Surprisingly, increasing incidents of myocardial injury with sustained cardiac involvement were found in 26-78% patients months after recovery from COVID-19^3,4^, with a proportion of patients with no known history of previous cardiac symptoms. This alarming prognosis suggests that cardiovascular complications are a long-lasting symptom of post-COVID conditions^5^, demanding additional research focus on cardiac involvement and cellular responses for clues to early prediction and prevention of COVID aftermath.

SARS-CoV-2 infection directly triggers respiratory illness with symptoms such as difficulty breathing, causing acute or chronic pulmonary and cardiovascular hypoxia^6-8^. Pathophysiological hypoxia may act at both systemic and cellular levels with severe cases of COVID-19 that often progress to acute respiratory distress syndrome (ARDS) and increased mortality^9^. Under low oxygen conditions as seen in COVID-19, hypoxia-inducible factor 1 alpha (HIF1α), the master regulator of hypoxia, is protected from oxygen-dependent proteolysis and accumulates in the nucleus, where it dimerizes with HIF1β (ARNT) and binds to hypoxia-response elements (HREs) to activate hypoxia-regulated signaling. HIF1α stabilization leads to an avalanche of transcriptional activities involving angiogenesis, proliferation, homeostasis, inflammation, and metabolic switch^6,9-11^. In mammalian hearts, HIF1α is essential for cardioprotection in pressure overload and ischemic preconditioning models, as well as in adaptation of oxygen supply with coordinated contractility and metabolic switch^12-15^. Recent studies suggested that stabilized HIF1α may markedly decrease ACE2 expression, the entry path of SARS-CoV-2, thus compromising viral entry to mitigate COVID-19 conditions^9,16-18^. Congruently, another study reported decreased pathogenicity of SARS-CoV-2 in high-altitude environments, possibly benefiting from reduced ACE2 levels in lungs of local inhabitants who are acclimatized to hypoxic conditions^19^. Together, this evidence suggest that cellular hypoxia is a direct and logical pathological response upon SARS-CoV-2 infection.

In this study, we examined HIF1α expression retrospectively in two groups of COVID-19 patients, those with preserved cardiac function with Ejection Fraction (EF) > 50% (preserved EF), and those with moderate to severe cardiac dysfunction with EF < 45% (low EF), using post-mortem heart autopsies with matching cardiac echocardiography records. In preserved EF hearts, HIF1α was found to be predominately expressed in non-myocytes, protecting endothelial cells against apoptosis. In low EF hearts, HIF1α can accumulate in cardiomyocyte nuclei and retain the nuclear envelope. Additionally, cardiomyocytes in low EF hearts show signs of sarcomere and myofibril abnormalities, consistent with severe cardiac function loss. These data suggest that cell-type dependent HIF1α expression is increased in hearts of COVID-19 patients with preserved cardiac function. Our approach not only utilizes the critical connection between clinical and histological data, but also establishes a direct link of cardiac cellular responses in hypoxia induced by SARS-CoV-2 infection with matching functional and histologic data, serving as guidance for potential therapeutic approaches in COVID-19 treatment.

## METHODS

### Patients

All studies and protocols were reviewed and exempted by IRB at Mount Sinai Hospital to be ‘not human subjects’. COVID-19 patients and non-COVID donors included in this study are listed in Table 1. Clinical echocardiography data were obtained from Mount Sinai Heart between March – December, 2020. Patients’ echocardiography data including both systolic and diastolic function were examined. All gender and age groups were considered as long as cardiac involvement meets the preserved (EF > 50%, n=4) or moderate to severe (EF < 45%, n=4) criteria with confirmed SARS-CoV-2 infection. Post-mortem histology samples of the selected patients were obtained from Mount Sinai Pathology. Healthy heart samples of patients with non-cardiac causes of death and no known COVID-19 history were obtained from Advanced Tissue Service (ATS) or were used as non-COVID controls (n=3). For TEM control, a formalin-fixed unembedded heart sample with no known COVID-19 history was obtained from Mount Sinai Pathology (n=1).

**Table 1.** EF groups of patients and healthy donors.

| Patient / Donor Groups | EF | Age Range | Gender | BMI | BSA | Brief disease condition |
| --- | --- | --- | --- | --- | --- | --- |
| **COVID-19, Preserved EF (> 50%, n = 4)** |  |  |  |  |  |  |
| N1 | 63% | 66-70 | M | 22.65 | 1.79 | diverticulitis, horseshoe kidney with no renal failure |
| N2 | 59% | 46-50 | F | 38.36 | 2.34 | Severely dilated RV / severe RV dysfunction |
| N3 | 58% | 71-75 | M | 24.01 | 1.95 | new ex lymphoma |
| N4 | 58% | 56-60 | F | 55.20 | 2.68 | Diabetes |
| **COVID-19, Low EF (<45%, n = 4)** |  |  |  |  |  |  |
| L1 | 43% | 71-75 | M | 47.00 | 2.54 | No pre-existing cardiac disease |
| L2 | 37% | 51-55 | M | na | na | CAD s/p MI in 2014 |
| L3 | 35% | 76-80 | M | na | na | CAD s/p CABG, HFrEF |
| L4 | 33% | 36-40 | M | 50.60 | 2.67 | PCA occlusion / thrombectomy / severe LV dialation |
| **Non-COVID-19, Healthy (n = 4)** |  |  |  |  |  |  |
| H1 | na | 56-60 | F | na | na | Unknown |
| H2 | na | 56-60 | M | na | na | Unknown |
| H3 | na | 76-80 | M | na | na | Unknown |
| H4 (TEM only) | na | na | na | na | na | Unknown |

### Immunostaining

Formalin-fixed paraffin-embedded heart sections were deparaffinized by serial immersion of Xylene, Absolute Ethanol, 95% Ethanol, 70% Ethanol. Antigen retrieval was achieved by microwaving slides in Citrus Buffer (pH 6.0) immersion at near boiling point for 12-15 min. Endogenous peroxidase activity was quenched by incubating in 3% H2O2 for 20min at room temperature. Following 1hr incubation in blocking reagent (APExBIO, K1051) with 10% Donkey Serum (Jackson Immunoresearch, #017-000-121), samples were incubated overnight in primary antibodies at 4ºC, washed in 1X TBST (Fisher, BP24711) with 0.05% Tween-20, and incubated in secondary antibodies (1:200) for 90min at RT. Antibodies used in this study: HIF1α (Abcam ab51608, 1:200), MyBPC3 (ThermoFisher PA547925, 1:50), Vimentin (Invitrogen PA1-16759), CTNT (Santa Cruz CT3 sc-20025, 1:100), CD31 (Santa Cruz M-20, sc-1506), Lamin B1 (SYSY HS-404017, 1:100), 8-Oxo-G (Abcam, ab62623, 1:100), Sars-CoV-2 Spike protein (GeneTex, GTX632604, 1:50). Spike protein staining was amplified using anti-mouse HRP (Invitrogen, SA1-100, 1:200) and Cy3 TSA fluorescence system kit (APExBIO, K1051). Autofluorescence background was quenched in 0.25% Sudan Black B (Sigma-Aldrich S-2380, in 70% Ethanol) for 5min at RT. Nuclei were counterstained with DAPI (ThermoFisher 62247, 0.1µg/mL), Sytox Blue (ThermoFisher S11348), or DRAQ5 (ThermoFisher 62251) for 15min at RT. Slides were then mounted in Mounting media (Vector Labs H-1000, or KPL 71-00-16) before imaging by Leica SP8 confocal microscopy.

### Apoptosis

TUNEL assay was performed using *in situ* cell death detection Kit (Roche 11684795910). Briefly, heart sections were deparaffinized and re-hydrated. Following brief wash in TBST, samples were incubated in TUNEL reaction mixture (Label solution:Enzyme solution, 9:1) for 1 hour at 37ºC HybEZ II Hybridization Oven (ACDbio). Slides were then washed in 1X TBST, blocked in blocking reagent, and continued with subsequent immunostaining.

### Quantitative RT-PCR

Total RNA was extracted from post-mortem lungs using RNeasy FFPE kit (Qiagen, 73504) and quantified using Qubit 3.0 Fluorometer. First strand cDNA was synthesized using High-Capacity cDNA reverse transcription kit (Applied Biosystems, 4368814). For Real-Time PCR reaction, 6.5ng cDNA was loaded in each reaction with SYBR Green PCR Master Mix (Applied Biosystems, 4309155) on an Applied Biosystems OneStep Plus thermocycler. Primers sequences used in this study are (5’-3’): *ACTB* Fw-GATTCCTATGTGGGCGACGA, Rv-AGGTCTCAAACATGATCTGGGT; *HIF1A* Fw-ATCCATGTGACCATGAGGAAATG, Rv-TCGGCTAGTTAGGGTACACTTC; *HIF1B(ARNT)* Fw-CTGCCAACCCCGAAATGACAT, Rv-CGCCGCTTAATAGCCCTCTG; *HIF2A(EPAS1)* Fw-GGACTTACACAGGTGGAGCTA, Rv-TCTCACGAATCTCCTCATGGT. Ct values were normalized to *ACTB* and analyzed by the ΔΔCt method relative to non-COVID controls.

### Transmission Electron Microscopy (TEM)

Samples were post-fixed in 2% paraformaldehyde and 2% glutaraldehyde in 0.1M sodium cacodylate buffer for 1 week, washed in several rinses of 0.1 M Sodium Cacodylate buffer, pH 7.2, followed by ddH2O rinses. Samples were further post-fixed with 1% aqueous osmium tetroxide, pH 7.2 for 1hr, and then En bloc stained in 2% aqueous uranyl acetate for 1 hr. Sections were washed again in ddH2O, dehydrated through graduated ethanol (25-100%) to propylene oxide series, and resin-infiltrated with an epoxy resin mixture (Electron Microscopy Sciences, EMS). Inverted #00 BEEM capsules (EMS) was used to accommodate for flat embedding of the material. These capsules polymerized in a vacuum oven at 60 °C for 72 hr. Ultrathin sections (85 nm) were sectioned with a diamond knife (Diatome) on a ultramicrotome (Leica UCT ultramicrotome) and mounted on 400 hexagonal mesh copper grids (EMS) using a Coat-Quick adhesive pen (EMS). Sections were counterstained with Uranyl Acetate and Lead Citrate. Sections were imaged on a Hitachi 7700 Electron Microscope and photographed with an Advantage CCD camera (Advanced Microscopy Techniques). Image brightness, contrast, and size were adjusted using Adobe Photoshop CS4 software.

Due to sample scarcity, formalin fixed, unembedded heart autopsies were used to prepare TEM (n = 1 of each group and control). Notice formalin may impair downstream TEM quality. Non-COVID control is a pathological sample with unknown prior cardiac history.

### Statistical Analysis

All data were quantified by ImageJ (analyze particle). Statistical analysis was performed using GraphPad Prism 9.0 with unpaired t test, nonparametric, Kolmogorov-Smirnov. *p* value < 0.05 was considered statistically significant.

## RESULTS

### COVID-19 patients and non-COVID controls

A total of 8 COVID-19 patients with random gender and age (range 39-76) were retrospectively selected and divided into two groups based on their left ventricular systolic function Ejection Fraction (EF). Patients with EF > 50% were categorized as “preserved EF” (range 58%-63%), and patients with EF < 45% were categorized as “low EF” (range 33%-43%). Two males and two females were included in the preserved EF group, whereas four males were included in the low EF group. Three non-COVID donor hearts were included as controls (age range 59-79). The median age of COVID-19 patients included in this study was 64, and the median age of non-COVID healthy donors was 60. Patient data and brief pre-existing cardiac conditions prior to COVID-19 diagnosis are listed in Table 1. Immunostaining of spike protein of SARS-CoV-2 confirmed the virus presence in patient lungs, whereas massive DNA lesions were also observed (Fig. S1).

### Cardiac HIF1α expression appears increased with preserved EF

To examine if COVID-19 patient tissues display any cellular signs of hypoxia, immunostaining of HIF1α was performed on lung sections, in which HIF1α expression was found to be significantly higher in patients with low EF in comparison to preserved EF group (Fig. S2A-C). Real-Time PCR revealed no apparent difference of HIF1α (*HIF1A)* and its dimerization units HIF2α (*EPAS1)* and HIF1β (*ARNT*) at transcript level in both groups (Fig. S2D-E). However, there is an inverse correlation in the heart. The number of HIF1α^+^ cells was significantly lower in low EF hearts than found in the preserved EF group (Fig. 1A-B, Fig. S3A-F). In non-COVID controls, very few to no HIF1α^+^ cells were observed (Fig. 1C, Fig. S3G-H). These results reveal a remarkable positive relationship between HIF1α expression and EF in COVID-19 patients (Fig. 1D, Fig. S3I), that HIF1α expression is upregulated in hearts with preserved function, indicating a protective role of HIF1α in preserving EF levels.

**Figure 1.**
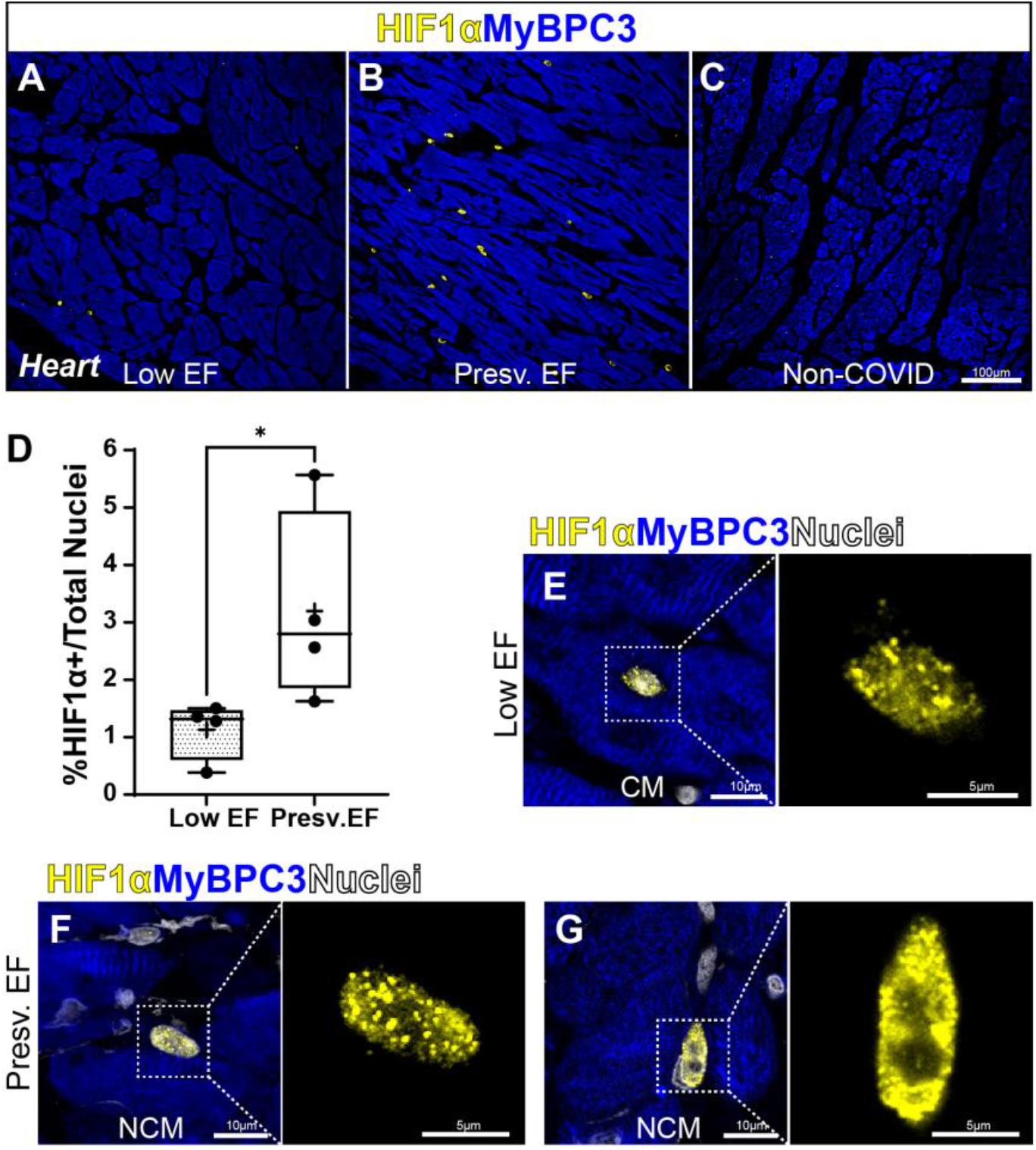
Cardiac HIF1α expression increases with EF in COVID-19 patients. Immunostaining showing HIF1α+ cells in COVID-19 patient left ventricles with A) lower EF (<45%, L4) than B) EF (>50%, N1). C) HIF1α expression is low to undetectable in non-COVID donor hearts (H3). D) Quantification of percentage of HIFα+ cells over total cell counted indicates a clear increase in number of HIF1α+ cells with higher EF%. Total nuclei counted: Low EF = 18,826 from n = 4 patients; presv. EF = 29,313 from n =4 patients. Data are represented as min to max plot from 3-4 independent experiments. Data range: Low EF=0.3841%-1.3585%, Presv.EF=1.6238%-5.5671%. Bar, median. +, mean. \**p* = 0.0286. Unpaired t test, nonparametric, Kolmogorov-Smirnov. Also see graph of individual patient in Fig. S3E. In low EF hearts, HIF1α is primarily expressed in cardiomyocyte nuclei. F-G) In Preserved EF hearts, HIF1α is primarily expressed in non-myocytes at F) nuclear or G) peri-nuclear localization. HIF1α, yellow. Myocyte, MyBPC3, blue. Nuclei, Sytox Blue, white. Scale bar, A-C: 100μm, E-G: 10μm and 5μm. Representative images are from n = 4 patients per EF group from 4 independent experiments.

In normoxia, HIF1α hydroxylation is catalyzed by von Hippel-Lindau (VHL) and shuttled by prolyl-hydroxylase domain proteins (PHDs) for ubiquitination and subsequent proteolysis^20-22^, both activities are oxygen-dependent. In hypoxia, this proteolysis ceases as HIF1α shuttles into the nucleus for its heterodimerization with HIF1β and DNA binding to HRE sequences, resulting in HIF1α nuclear accumulation. In COVID-19 hearts, HIF1α primarily accumulated into speckles in nuclei of cardiomyocytes (CM) in low EF patients (Fig. 1E). To the contrary, HIF1α was predominantly detected only in non-myocytes (NCM) with either nuclear or peri-nuclear localization in preserved EF patients (Fig. 1F-G). This cell-type dependent differential expression pattern in low and preserved EF tissues may indicate distinct roles of HIF1α in different cell types.

### HIF1α is protective against apoptosis in non-myocytes

Cardiac non-myocytes consist of a heterogeneous collection of cell types. Among these, fibroblast and endothelial cells comprise the largest cell distributions, representing almost 60% cells in adult human atrial and ventricular tissues^23^. Therefore, we next investigated the hypoxic response in fibroblasts and endothelial cells in COVID-19 hearts.

Abundant accumulation of HIF1α was detected in the pulmonary valve (PV) in comparison to myocardium (Fig. 2A, 1 and 2), indicating that valvular tissue is more prone to hypoxic distress. Co-expression of Vimentin and HIF1α in the valve further demonstrates that cardiac fibroblasts can be susceptible to hypoxia (Fig. 2B).

**Figure 2.**
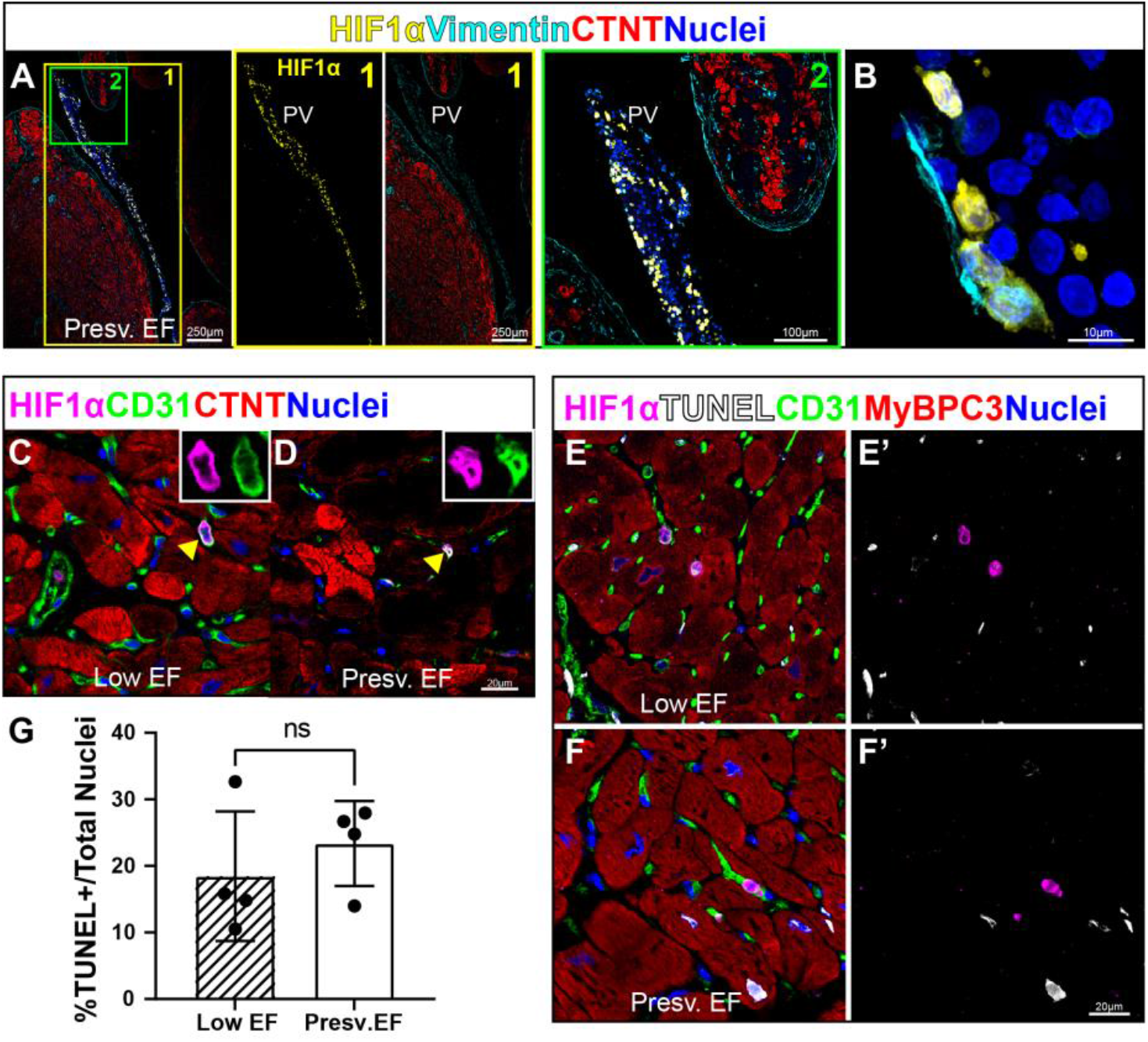
HIF1α expression in endothelial cells and cardiac fibroblast is cardioprotective against apoptosis. A) Abundant HIF1α cells in RV/IVS valve region. B) co-expression of HIF1α and Vimentin indicating fibroblasts can also be prone to hypoxia in a Presv. EF heart. HIF1α, yellow. Vimentin, cyan. Myocyte, CTNT, red. Nuclei blue. PV, pulmonary valve. C-D) Co-expression of HIF1α and CD31 showing endothelial cells are prone to hypoxia in both low EF and Presv. EF hearts. E-F) HIF1α+ are TUNEL-in both low EF and Presv. EF hearts. HIF1α, magenta. CD31, green. Myocyte, CTNT or MyBPC3, red. Nuclei, blue, TUNEL, white. Scale bar: A, 1: 250μm, 2: 100μm, B: 10μm, C-F: 20μm. G) Quantification of TUNEL+ cells in Low and Presv. EF showing no significant difference of apoptosis between the two groups. Total nuclei counted: Low EF = 8,853 from n = 4 patients; presv. EF = 15,984 from n =4 patients. Unpaired t test, nonparametric, Kolmogorov-Smirnov. Data are represented as Mean ± SD from 4 independent experiments.

Endothelial damage can be a direct consequence of SARS-CoV-2 infection due to expression of ACE2 receptor in endothelial cells^11,24^. Endothelial cells adapt to hypoxia by activating HIF1α and orchestrating a number of genes involving cellular metabolism, anti-apoptosis, and pro-inflammatory response^14,25^. In COVID-19 ischemic hearts, hypoxia is expected to play an essential role modulating endothelial cell survival. As expected, HIF1α was found in CD31^+^ endothelial cells in both low and preserved EF groups (Fig. 2C-D). Notably, all HIF1α^+^ cells in the heart were TUNEL negative (Fig. 2E-F). In lung, HIF1α expression also coincides with surviving cells whereas very few HIF1α^+^ cells being apoptotic (Fig. S4A-B, arrowhead showing a HIF1α^+^ /TUNEL^+^ double positive cell). This data revealed that HIF1α^+^ cells are resistant to apoptosis in both heart and lung, despite the total count of apoptotic cells appearing to be similar between low and preserved EF groups (Fig. 2G, Fig. S4C). These findings further support our hypothesis that HIF1α stabilization is a protective mechanism of COVID-19 hearts by preventing cell death of non-myocytes, specifically, by mediating endothelial cell survival under low oxygen conditions.

### Nuclear accumulation of HIF1α^+^ maintains cardiomyocyte nuclear stability

In the heart, cardiomyocytes are the only cell type reported to accumulate HIF1α in the nucleus during mouse cardiogenesis^26^, suggesting that nuclear accumulation of HIF1α under hypoxia may hold relevance to protecting ischemic myocardium in severe COVID-19 conditions. To test this hypothesis, we first examined nuclear and sarcomeric ultrastructure by transmission electron microscopy (TEM). In the preserved EF heart, the nuclear envelope appears to be thick and dense, similar to the non-COVID control. In the low EF heart, the nuclear envelope is significantly thinner, yet is maintained in a continuous pattern with no apparent breaks in the lamin membrane (Fig. 3A-C). Consistent with severe function loss, signs of sarcomeric damage are seen in the low EF heart as evident by the swollen Z lines, smeared I bands, shortened sarcomeres and distorted myofibril arrangements, whereas such structures are maintained in the preserved EF group as compared to non-COVID control (Fig. 3D-F). Fewer mitochondria were found in both COVID-19 groups than in control hearts. They scattered among myofibrils, rather than being regularly aligned in rows as seen in the control. Additionally, mitochondria in the low EF group appear to be either shrunk or swollen as signs of dysfunction under stress, consistent with cardiac function. This observation provides valuable structural information in explaining the difference in EF seen in each group.

**Figure 3.**
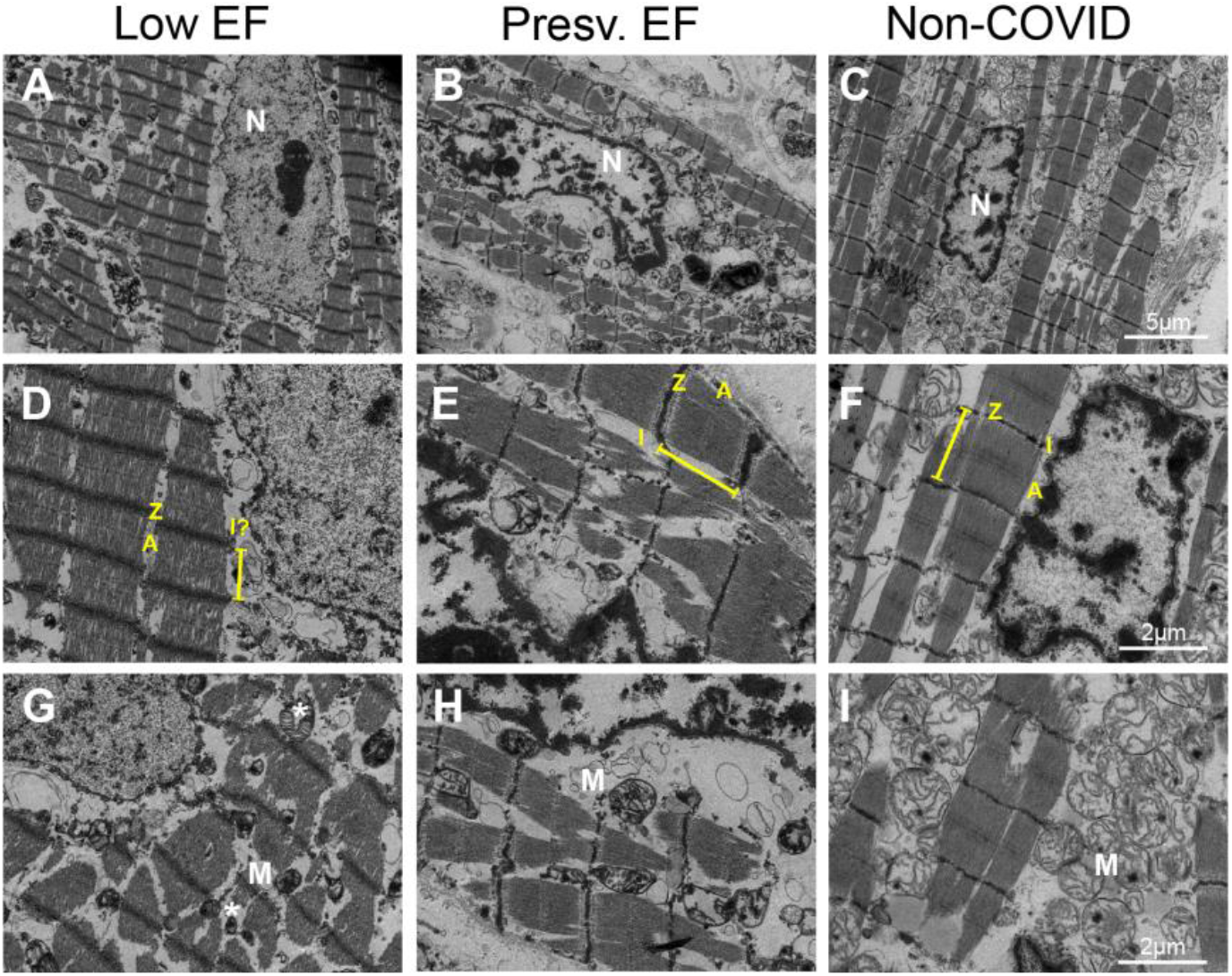
TEM of Preserved EF and Low EF hearts. TEM of COVID-19 hearts and non-COVID control showing striking difference of A-C) Nuclear ultrastructure, D-F) sarcomere structure and myofibril arrangement, and G-I) mitochondria, between Presv. EF and Low EF groups. N, nucleus. Z, z line. A, A band, I, I band. M, mitochondria. Asterisk, shrunk or enlarged mitochondria. A-C) low power. Scale bar, 5μm. D-I) high power. Scale bar, 2μm. Non-COVID control was obtained from a pathological autopsy with no known SARS-CoV-2 infection or cardiac condition. Due to sample scarcity, representative images are n = 1 heart per group (5-8 scans per heart).

It should be noted that cellular HIF1α expression level remain unknown in TEM. Therefore, we next examined heart sections by co-immunostaining with nuclear envelope marker Lamin B1 and HIF1α. In non-COVID hearts, Lamin B1 is continuously expressed in both cardiomyocytes and non-myocytes, with no significant difference between the two cell types (Fig. 4A). In COVID-19 cardiomyocytes that lack HIF1α expression, Lamin B1 expression is weakened but remains visible in the preserved EF group (Fig. 4B), whereas it becomes undetectable in the low EF group (Fig. 4C). Nuclei of these HIF1α^-^ myocytes appeared to be enlarged or irregularly shaped, a “leaky” phenotype that indicates unhealthy nuclei (Fig. 4B-C). This observation is in line with a recent report of loss of nuclear DNA in SARS-CoV-2 infected iPS-derived cardiomyocytes *in vitro*^2^. Surprisingly, Lamin B1 was found to be preserved normally in cardiomyocytes with nuclear HIF1α accumulation (Fig. 4D, also see Fig. S5 for more biological replicates), with a healthy spindle-shaped nuclear morphology. Together, this evidence strongly supports a protective role of HIF1α in maintaining DNA stability in cardiomyocytes.

**Figure 4.**
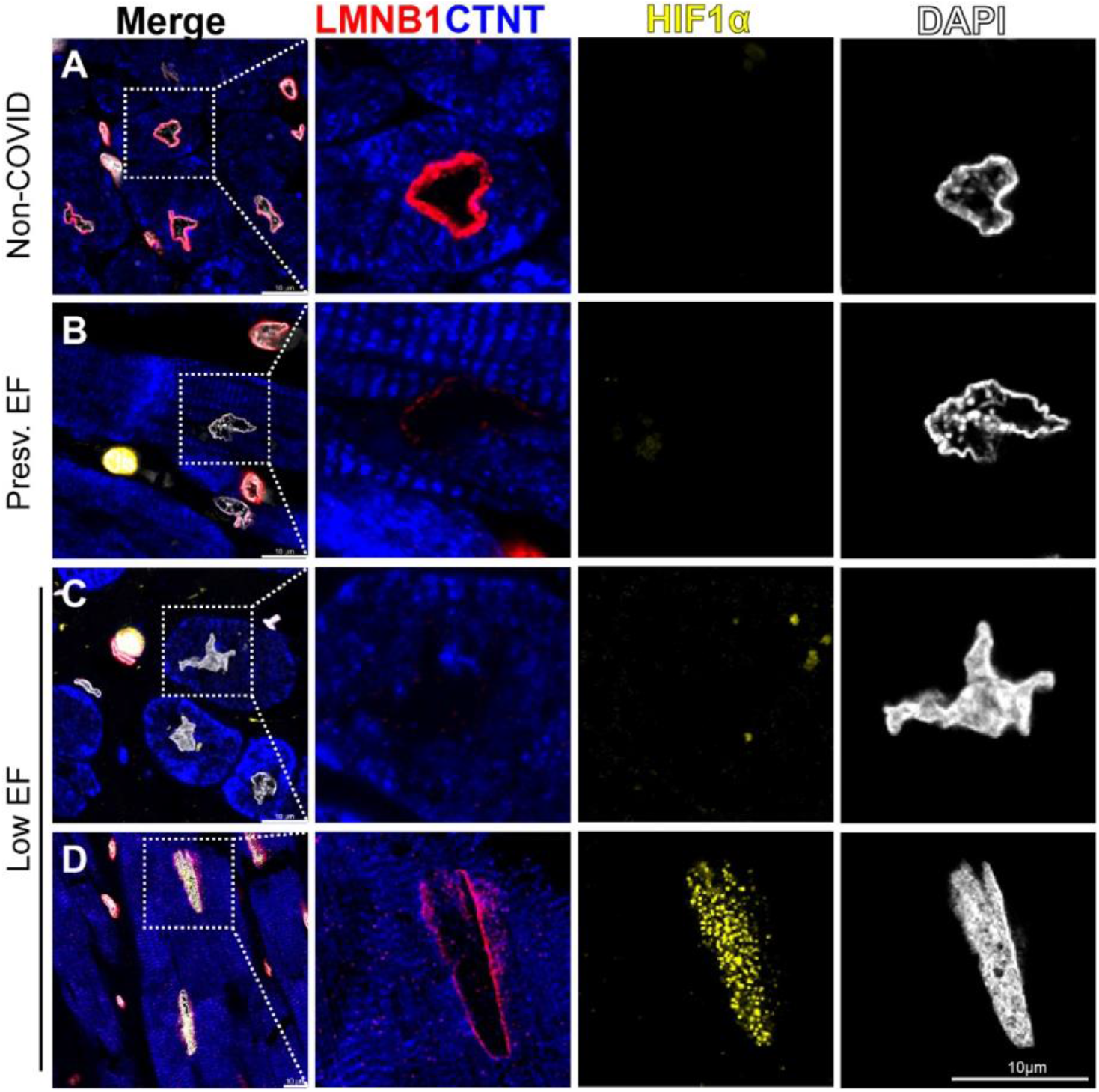
Nuclear accumulation of HIF1α maintains cardiomyocyte nuclear stability in COVID-19 hearts. A) Immunostaining of Lamin B1 in non-COVID donor hearts showing intact nuclear envelope in healthy myocytes (n = 3). B) A HIF1α-neg presv. EF myocyte with dim Lamin B1 expression and leaky nuclear morphology. C) A HIF1α-neg low EF myocyte with almost undetectable Lamin B1 and leaky nuclear shape. D) A low EF myocyte with nuclear HIF1α accumulation showing grossly intact Lamin B1 and compact nucleus morphology. Boxed area in left column (Merge) are magnified in separate channels. LMNB1, red. CTNT, blue. HIF1α, yellow. Nuclei, DAPI, blue. Scale bar, 10µm. Also see Fig. S6 for more biological replicates. Representative images are from n = 4 patients per EF group from 3 independent experiments.

## DISCUSSION

In this study, we report a positive relationship of HIF1α expression and cardiac function in COVID-19 patients, whereby HIF1α appears to protect endothelial cells against cell death and maintains cardiomyocyte nuclear integrity (Fig. 5). Cell-type specific HIF1α expression and subcellular localization strongly suggest a protective role of HIF1α in hypoxic cardiac environments.

**Figure 5.**
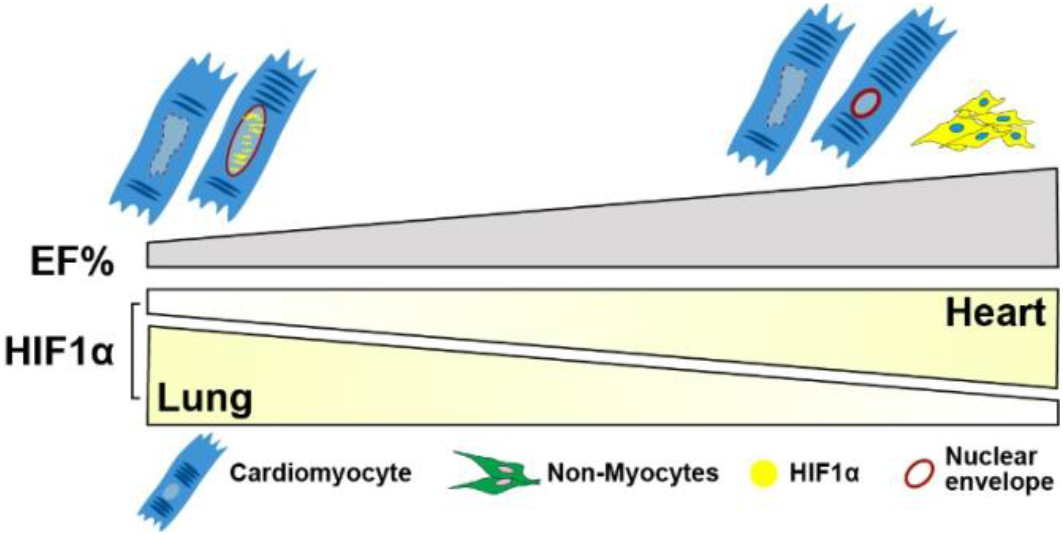
Schematic of HIF1α levels versus EF% in lung and heart. HIF1α expression is decreased in the lungs of COVID-19 patients with preserved cardiac EF but inversely, noted to be increased in the hearts of the same patients. In the preserved EF group (EF 58-63%), more HIF1α cells were detected predominantly in NCM cytoplasm (yellow shade). In low EF group (EF 33-43%), overall HIF1α^+^ cell quantity is lower whereas most HIF1α expression is detected in CM nuclei (yellow speckles). In cardiomyocytes, nuclear HIF1α appears to be protective by preserving nuclear Lamin and morphology. Myocyte nuclei that lack HIF1α expression appear to be enlarged and leaky with undetectable nuclear envelopes.

Post-mortem histology analysis of cellular damage holds critical clues to explain cardiac functional loss. However, such an important connection is largely missing as clinical cases and laboratory studies are often reported separately. For example, in mildly affected COVID-19 patients, cardiac functional data could be overlooked and recovered patients could continue to suffer from chronic cardiovascular abnormalities as reported in SARS-CoV virus cases^5,6,8^. On the other hand, severe cardiomyocyte damage has been reported in COVID-19 patients including necrosis, z-band rupture, and entire nuclear DNA loss^2,27,28^, but very limited cardiac functional data was reported to supplement the cellular findings. Though cardiomyocyte damage is significant, it is impossible to correlate such phenotypes with cardiac involvement without functional data, rendering an incomplete understanding of COVID-19 cardiac symptoms. With the advantage of direct accessibility of clinical echocardiography data and cardiac tissues obtained at autopsy from the same patients at Mount Sinai, we were able to fill the gap by carrying out our investigation in tissues with matching cardiac functional records. Therefore, cellular findings in this study can be directly associated with functional loss and cardiac involvement for comparison purposes.

A variety of roles and mechanisms of the HIF1α pathway have been reported in recent COVID-19 studies. In lung, alveolar type II (AT2) cells present ACE2 receptors and are a direct entry path of viral infection, causing inflammation and hypoxia^6^. It has been speculated that HIF1α drives pro-inflammatory cytokines that lead to cytokine storm in severe COVID-19 cases^6^. Contrarily, this hypoxia-mediated cell inflammation can proportionately downregulate CD55 and protect complement-mediated endothelial damage, assist glycolytic metabolic switch and permit ATP generation under hypoxic conditions, ultimately preventing apoptosis^25^. Indeed, our data captured such distinct HIF1α expression in lungs and hearts of the same patients. Patients with compromised EF showed high HIF1α expression in lung but decreased HIF1α expression in heart, indicating that HIF1α may play opposite roles in lung and heart. This evidence also argues that cellular responses of SARS-CoV-2 infection can be overlooked on a systemic level and should be investigated in a tissue-specific manner.

In cancer treatment, hypoxic microenvironments can protect tumor cells from apoptotic stimuli *in vivo*^29^. Low-oxygen regions constantly select cells with enhanced ability to withstand treatment and lead to increase resistance to apoptosis^30^. Surprisingly, this is consistent with our findings that hypoxic cells were resistant to apoptosis in COVID-19 hearts independent of cardiac involvement, as surviving HIF1α^+^ endothelial cells were observed in both low and preserved EF groups. It is remarkable how damaged endothelium thrives to adapt to a low oxygen environment after SARS-CoV-2 infection. As such, HIF1α pathway may hold promise as a potential therapeutic target. Further investigation of HIF1α pharmacological reagents in endothelial cells, vessel barrier integrity, as well as vascular inflammation will provide insights regarding their therapeutic value especially for COVID-19 patients with cardiovascular disease.

Our immunostaining consistently shows that HIF1α clusters into speckles when it accumulates in cardiomyocyte nuclei, a unique distribution pattern of endogenous HIF1α, HIF2α, and HIF1β previously seen in immortal cell lines^31^. Nuclear speckles are often associated with active transcription binding sites with nuclear protein and molecules trafficking at rapidly dynamic mobility. This nuclear distribution is consistent with a HIF1α transcriptional master controller role under low oxygen. Additionally, packaging into nuclear speckles may hinder HIF1α/HIF2α heterodimer mobility therefore protects HIF1α from cytosolic degradation.

Hypoxia stress is often linked with genetic instability and higher mutagenesis rate in tumor treatment, with possible escape of surveillance from a variety of transcription and translation factors such as DNA repair, cell-cycle checkpoint, and chromatin structure maintenance^32^. Our data shows that nuclear HIF1α^+^ cardiomyocytes are capable of preserving intact nuclear envelopes during COVID-19-mediated hypoxia, although the degree of genetic mutation or the duration of hypoxic exposure remain unknown. Further investigation on ploidy, DNA damage and surveillance are required to provide more insights into the mechanism behind HIF1α and genomic stability.

Our data provide compelling evidence of the protective role played by HIF1α in hearts of patients affected by COVID-19. We believe this is one of the first studies to bridge previous stand-alone clinical data and cellular data, with valuable evidence of cardiac cellular characterization after SARS-CoV-2 infection that encompasses a structure-function relationship in the same individual patients. We believe further study on this topic may shed insights into mechanistic pathways mediating other forms of viral cardiomyopathy.

## Data Availability

The data that support the findings of this study are available from the corresponding author upon reasonable request. All data generated or analyzed during this study are included in this published article (and its supplementary information files).

## ACKNOWLEDGEMENT

We gratefully acknowledge Mount Sinai Heart for providing COVID-19 cases in this study. Sincerely thanks to Frances Avila, Alan Soto, and Dr. Yayoi Kinoshita at Biorepository CoRE for their general facilitation with specimen preparation. EM tissue preparation and imaging was performed at the Microscopy CoRE and Advanced Bioimaging Center at Mount Sinai and we thank Allison Sowa and Bill Janssen for their timely assistance with TEM.

## AUTHOR CONTRIBUTIONS

B.J.W. and H.W.C. designed the overall experiments. R.I.B. and L.B.C. provided patient cases and histological samples for data collection. B.J.W., L.B.C., and S.V.M. performed the experiments. B.J.W. and H.W.C. analyzed the data. B.J.W. and H.W.C. wrote the manuscript. All authors read and approved the final manuscript.

## DATA AVAILABILITY

The data that support findings of this study are available from the corresponding author upon reasonable request. All data generated or analyzed during this study are included in this published article (and its supplementary information files).

## SUPPLEMENTAL FIGURES

**Figure S1.**
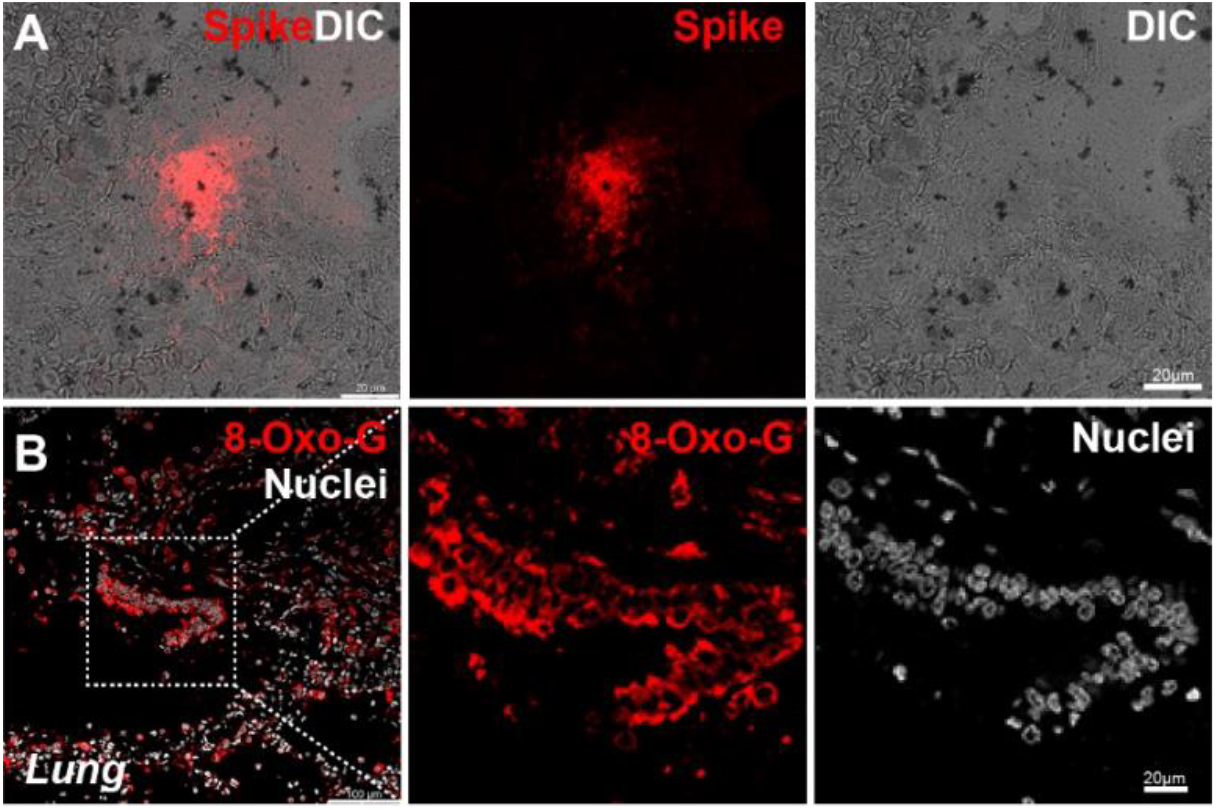
SARS-CoV-2 infection and damage in lungs. A) Immunostaining of Spike protein of SARS-CoV-2 confirming the presence of viral protein (Red) in patient lung (L1). DIC, differential interference contrast. B) Immunostaining of 8-Oxo-G (Red) showing lesion damage in lung (N4). Nuclei, DRAQ5 (White). Scale bar, 20µm. Representative images are from n = 2 patients from 3 independent experiments.

**Figure S2.**
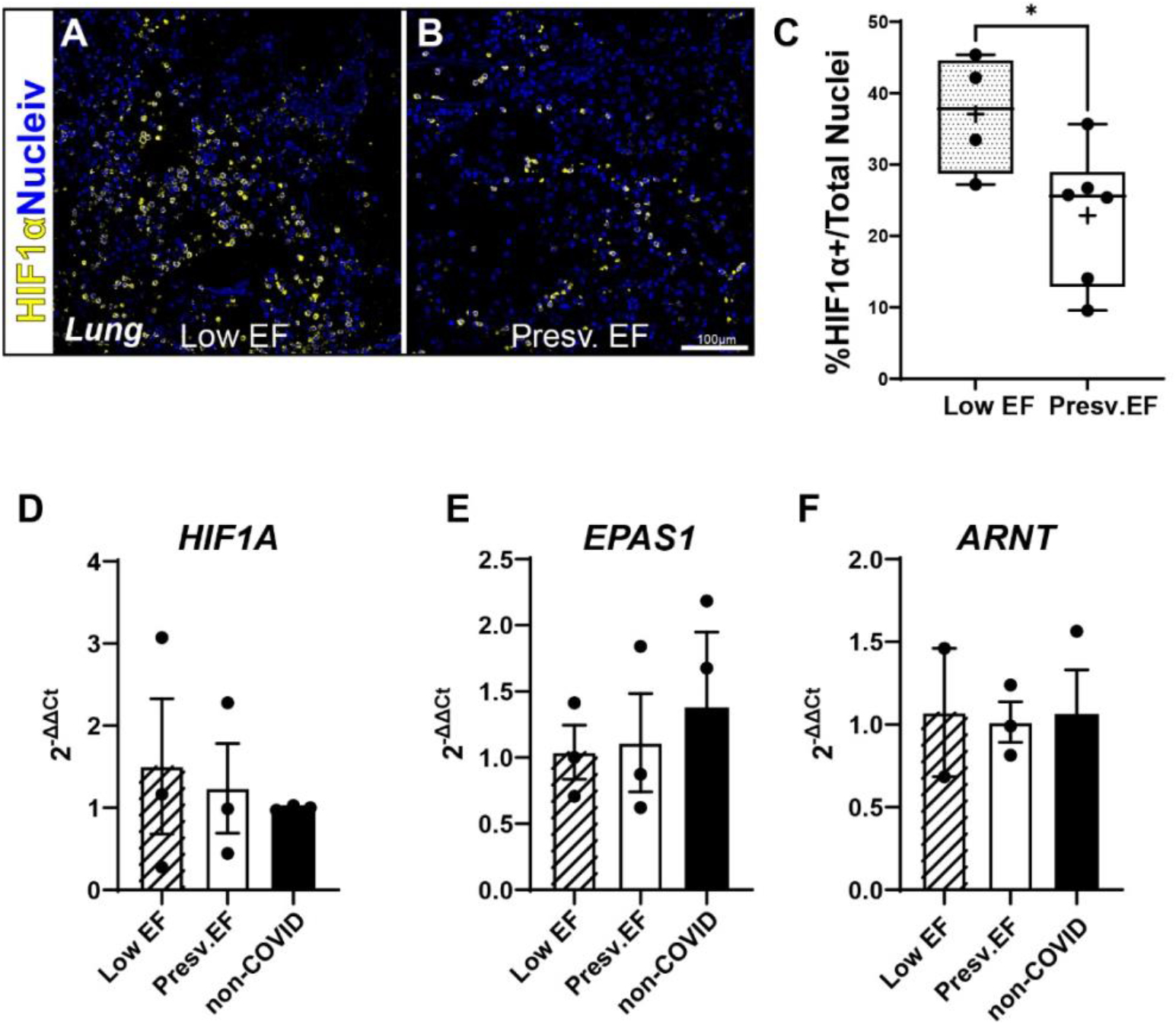
HIF1α expression in lungs. Immunostaining of HIF1α in lungs of COVID-19 patients with A) low EF (L1, EF=43%) and B) Presv. EF (N4, EF=58%). HIF1α, yellow. Nuclei, DAPI, Blue. Scale bar, 100µm. C) Quantification of percentage of HIFα+ cells over total cell counted indicates an inversed correlation of number of HIF1α+ cells vs. EF%. Total nuclei counted: Low EF = 8,170 from n = 1 patient due to sample scarcity; presv. EF = 10,437 from n =3 patients. Data are represented as min to max plot from 4-6 independent experiments. Low EF=27.2%-45.4%, Presv.EF=9.6%-35.7%. Bar, median. +, mean. *p = 0.0476. D-F) Expression of *HIF1A, EPAS1*, and *ARNT* showing no significant difference of HIF1α and its dimerization factors at transcript level in lungs of low and presv. EF groups. Unpaired t test, nonparametric, Kolmogorov-Smirnov. Quantifications are represented as mean±SEM from n = 3 per group from 4 replicates.

**Figure S3.**
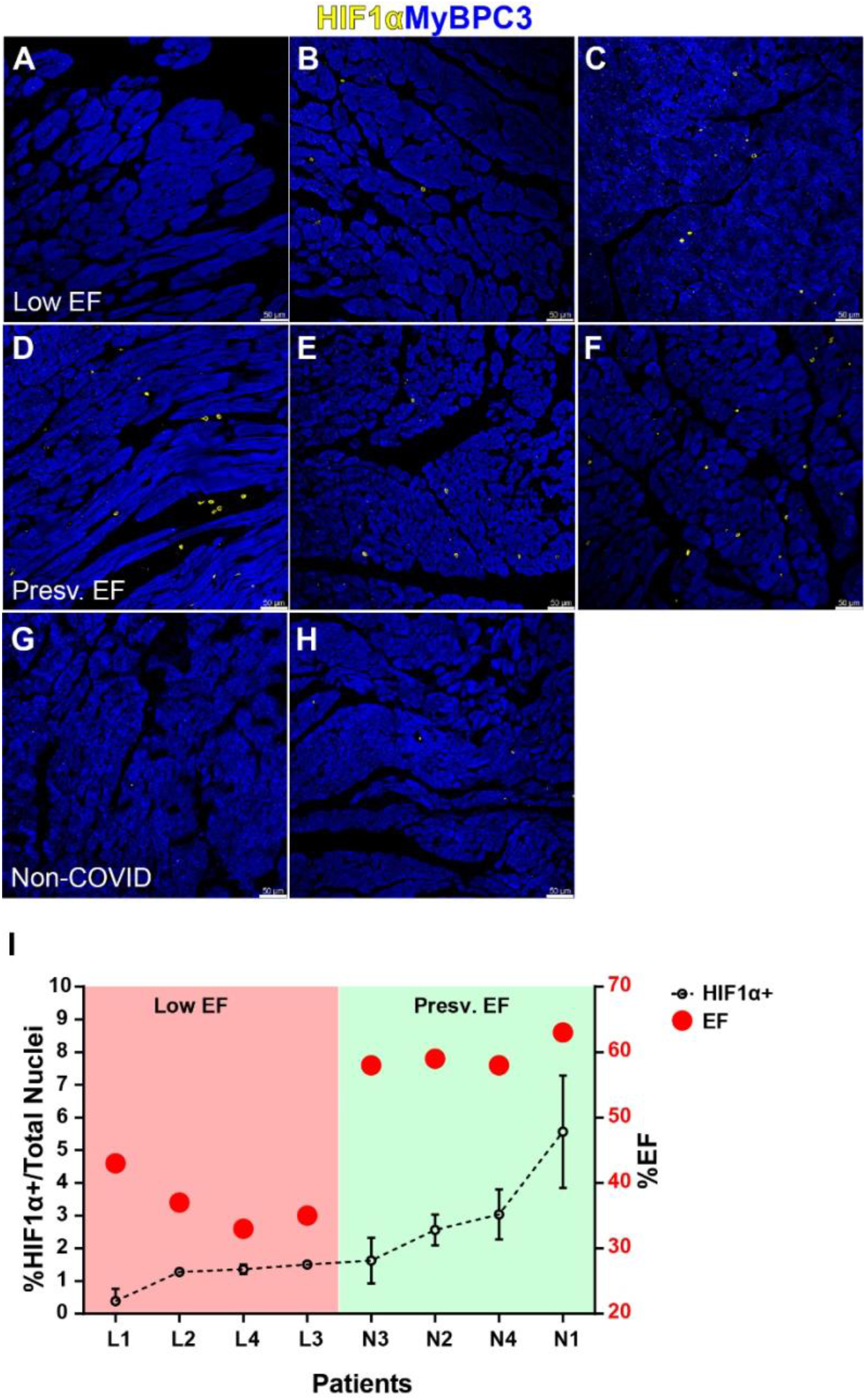
HIF1α expression in individual hearts. Representative images of HIF1α expression in each patient and donor heart included in this study. A-C) COVID-19 patients with low EF (L1, L2, L3) (also see Fig. 1, n = 4 total). D-F) COVID-19 patients with Presv. EF (N2, N3. N4) (also see Fig. 1, n = 4 total). G-H) non-COVID donors (H1, H2) (also see Fig. 1, n = 3 total). HIF1α, yellow. Myocyte, MyBPC3, blue. Scale bar, 50µm. I) Two-axis plot of HIF1α expression and EF% of each patient as an individual. Pooled group data is presented in Figure 1. Coral shaded area: Low EF group and less HIF1α expression, Lime shaded area: Presv. EF group with more HIF1α expression. Error bars respresent Mean±SD from 3-4 replicates per patient.

**Figure S4.**
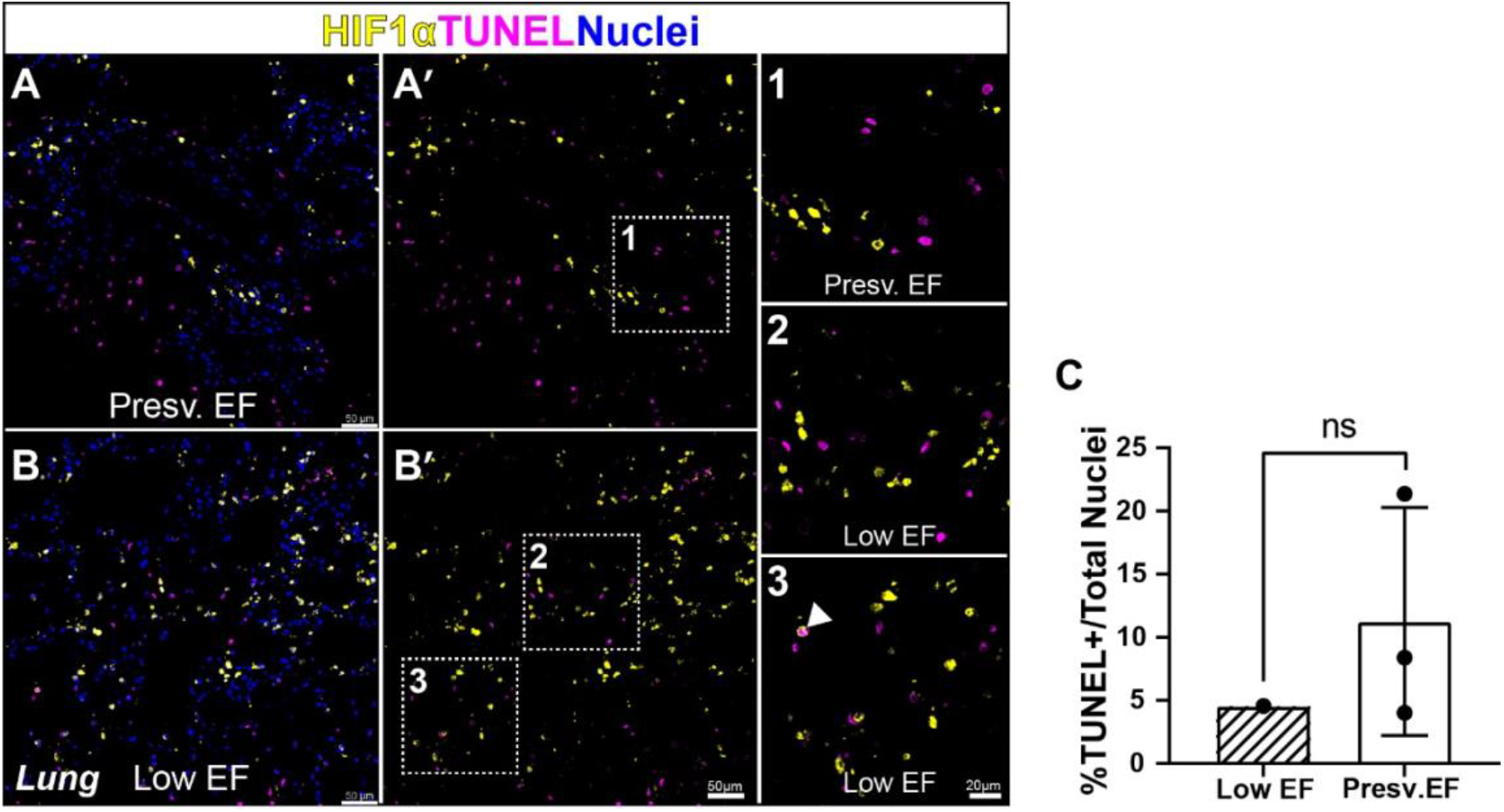
HIF1α and apoptosis in lungs. TUNEL and co-immunostaining of HIF1α+ showing are HIF1α+ cells are rarely apoptotic in COVID-19 lungs. A) in Presv. EF patients, HIF1α+ cells are TUNEL-. B) in low EF patients, vast majority of HIF1α+ cells are TUNEL-with very few apoptotic HIF1α+ cells detected (arrowhead). Boxed region is magnified in 1-3. HIF1α, yellow. TUNEL, magenta, Nuclei, blue. Scale bar, A-B: 50μm, 1-3: 20μm. C) Quantification of TUNEL+ cells in Low and Presv. EF showing no significant difference of apoptosis between the two groups. Total nuclei counted: Low EF = 6,282 from n = 1 patient due to sample scarcity; presv. EF = 6,313 from n = 3 patients. Unpaired t test, nonparametric, Kolmogorov-Smirnov. Data are represented as Mean±SD from 2 independent experiments.

**Figure S5.**
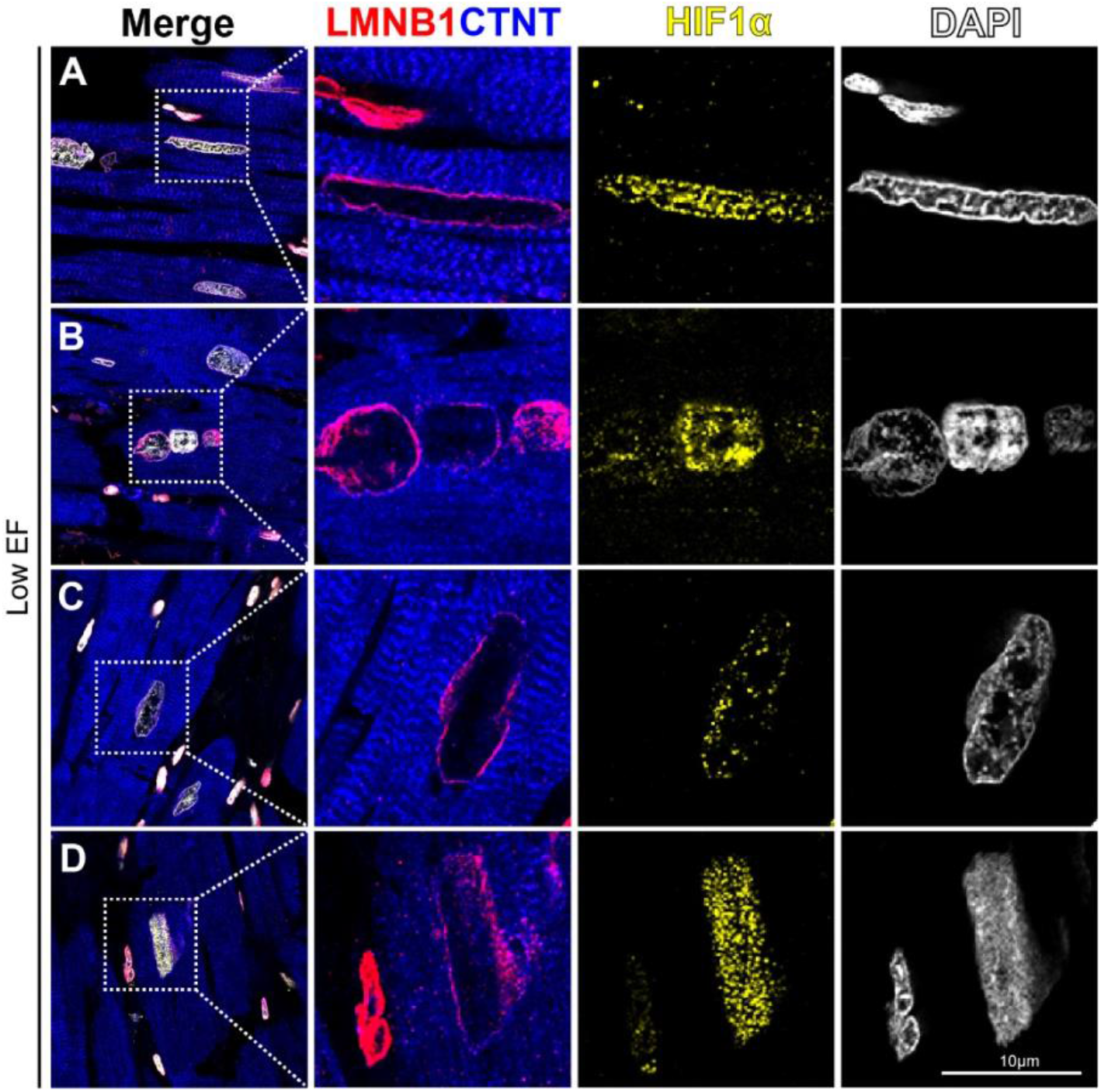
Representative images of preserved Lamin B1 in cardiomyocytes with nuclear HIF1α. Preserved nuclear envelope and compact nuclear morphology in myocyte with nuclear HIF1α accumulation low EF hearts. A) and B) L3, C) L1, D) L4. Boxes area in left column (Merge) are magnified in separate channels. LMNB1, red. CTNT, blue. HIF1α, yellow. Nuclei, DAPI, blue. Scale bar, 10µm. Representative images are from n = 4 patients per EF group from 3 independent experiments.

